# Design and Validation of a Pragmatic, Scalable Prioritization Tool for Cognitive Screening using the Electronic Health Record

**DOI:** 10.1101/2025.04.02.25325089

**Authors:** Byron C. Jaeger, Joseph Rigdon, Mat Weiss, Paul Yelton, Nori Allen, Nicholas M. Pajewski, Gabriel S Tajeu, Suzanne Craft, Michelle M. Mielke, Jeff D. Williamson

## Abstract

**INTRODUCTION:** Dementia is a disabling condition that progressively impairs daily function. Timely identification of older adults at high risk for dementia or cognitive impairment (a potential precursor) is critical to maximizing opportunities for intervention.

**METHODS:** Utilizing structured electronic health record data from 122,633 patients aged 55-80 years, we leveraged demographics, encounter diagnoses, and patient problem lists to develop and prospectively validate a ML model.

**RESULTS:** The ML model achieved a C-statistic of 0.811 (95% confidence interval: 0.810, 0.812) with adequate calibration overall and in subgroups based on race and sex. Recommending screening for patients with 3-year predicted risk > 5%, the ML model obtained satisfactory fairness across race and sex subgroups, with a net benefit of 18 true positive MCI/dementia diagnoses per 1,000 patients.

**DISCUSSION:** The ML model developed in this study can effectively identify individuals at high risk for a future diagnosis of MCI/dementia, potentially facilitating earlier screening and intervention to reduce the burden of cognition-related disability.

## INTRODUCTION

Dementia is a disabling condition affecting memory, thinking, and social abilities that progressively impairs daily function. Prior studies estimate over 9 million Americans could have dementia by 2030 and nearly 12 million by 2040.^1^ The number of adults with mild cognitive impairment (MCI), a potential precursor for dementia, is also high and expected to rise with the increasing age of the population.^1^ While the US Preventive Services Task Force currently does not recommend cognitive screening in older adults,^2^ the landscape is changing with the recent approval of disease-modifying therapeutics for Alzheimer’s disease and growing clinical trial evidence for the benefit of modifiable risk factors such as blood pressure control.^3–5^ Timely identification of older adults at high risk for incident cognitive impairment is critical to maximizing opportunities for intervention.

Machine learning (ML) algorithms have shown capacity to leverage biomedical and genetic data to identify people with high risk of developing numerous adverse health conditions, with several recent models targeting the prediction of MCI and/or dementia.^6,7^ However, many prior models have leveraged case-control designs (removing any ability to calibrate the model), have required unstructured data or specialized predictors that are not widely available in the electronic health record (EHR), have not accounted for timing and/or censoring of incident diagnoses of MCI and/or dementia, or require specialized and computationally complex processing. Few studies have evaluated the prognostic value of ML algorithms trained only using readily available information, including demographics, diagnoses, and medical conditions (i.e., problem lists) to predict future risk for incident diagnoses of MCI or dementia. Restricting ML algorithms to these data elements may improve scalability, as they do not require further data collection from patients or providers and are consistently available in most EHRs. We analyzed EHR data from Atrium Health-Wake Forest Baptist (AH-WFB), an academic health system in North Carolina, to evaluate whether ML algorithms leveraging these broadly available data elements can effectively and equitably identify who to recommend for cognitive screening based on the predicted risk of a future MCI or dementia diagnosis, with the downstream goal of identifying persons eligible for various preventive interventions.

## METHODS

### Study population

AH-WFB is a six-hospital healthcare system in the Piedmont region of the southeastern United States comprising a 24-county region including western North Carolina and southern Virginia.^8^ Primary care is organized through over 1,000 providers within an integrated network of over 165 practices. AH-WFB has used the Epic (Verona, WI) EHR system across its inpatient and outpatient settings since 2013.

We identified patients with at least one ambulatory office visit during a study period that spanned from 01/01/2019 to 07/01/2021. We restricted analyses to patients with at least one valid “index visit”, defined as an ambulatory office visit where the patient:

1. Was 55 to 80 years of age at the time of the office visit,
2. Had at least 1 outpatient visit prior to the index visit,
3. Had at least 1 outpatient visit after the index visit,
4. Had at least 365 days since their first outpatient encounter in the healthcare system
5. Had no prior diagnoses of MCI or dementia.

The age range of 55 to 80 years was selected to target a population that would be most likely to benefit from treatment to slow or reverse cognitive decline. For patients with multiple valid index visits, a single index visit was selected at random from the set of valid visits. The index date was defined as the date that the index visit occurred.

### Primary outcome

The primary outcome was time from the index date to an incident diagnosis of MCI, dementia, or the last recorded outpatient encounter, whichever occurred first. Diagnoses of MCI and dementia were ascertained by validated codes from encounter diagnoses or if they were indicated on the problem list. Incident dementia events were identified and classified according to ICD-10 codes (Table S1).

### Learners

We assessed five learners (i.e., methods to develop prediction models) including traditional and penalized Cox regression,^9^ axis-based and oblique random survival forests (RSFs),^10,11^ and boosted decision trees.^12^ A super learner was derived by combining weighted predictions from the individual learners.^13,14^ The weights are initialized as 1/L, where L is the number of learners, and then iteratively updated to maximize a chosen prediction accuracy metric.

### Predictors

Predictors were derived from patient data during a ‘look-back’ period that included outpatient visits occurring after 2015-01-01 and before or on the index date. A total of 10 conditions known to be associated with cognitive impairment were included: atrial fibrillation, cardiovascular disease, depression, diabetes, head trauma, hypertension, insomnia, orthostatic hypotension, stroke, and family history of Alzheimer’s Disease. The elapsed time from diagnosis and problem list indication to index date was derived for each condition, as was the relative frequency of the condition’s appearance as a diagnosis and as a problem on the problem list. In addition, we included age on the index date, sex, and race/ethnicity as predictors.

#### Predictor selection

Predictors were selected using recursive feature elimination by oblique RSFs.^15,16^ We fit an initial oblique RSF using all available predictors and recorded its out-of-bag prediction accuracy. The term “out-of-bag” refers to a set of observations that were not used while growing a particular tree in the RF, and aggregating out-of-bag predictions over all trees in the RF allows for efficient approximation of external prediction accuracy.^17^ We then repeatedly dropped the ‘least important’ predictor, re-trained the oblique RSF, and recorded out-of-bag prediction accuracy until all predictors were dropped. The importance of predictors was calculated by running analysis of variance tables at every split in each oblique decision tree, and tracking the proportion of times each variable obtained a p-value <0.01.^18^ The set of predictors that obtained the highest out-of-bag prediction accuracy was designated as the selected set of predictors.

#### Predictor sets

We assessed three starting predictor sets (i.e., initial sets of candidate predictors prior to our variable selection step): diagnosis codes only, problem lists only, and the combination of diagnosis codes and problem lists.

### Internal and external validation

A training and testing set were created based on patient index date. Patients with index dates from 01/01/2019 to 07/01/2021 were assigned to the training data, while patients with index dates from 07/02/2021 to 07/01/2022 were assigned to the testing data. The testing data was reserved for prospective temporal validation and interpretation of the ‘final’ prediction model, while the training data was first leveraged to conduct 10-fold cross validation (i.e., internal validation) and then to fit the final prediction model. All data processing steps were replicated during cross-validation (i.e., predictor selection and determination of coefficients for scaling and imputing predictors) to ensure model performance estimates were not overly optimistic.^19^

### Prediction evaluation and explanation

All predictions were evaluated at 3-years after the index date overall and in subgroups based on race and sex. As a total of 2571 Hispanic patients were included in the prospective validation set with 117 MCI/dementia diagnoses, we did not assess predictions in subgroups based on Hispanic ethnicity. We assessed discrimination, calibration, index of prediction accuracy (IPA), net benefit, fairness, sensitivity, specificity, and positive/negative predictive value.^20,21^ Discrimination was estimated by concordance (C-) statistic, which measures the probability of assigning higher risk to a case versus a non-case.^22^ Calibration was evaluated in terms of agreement between average observed risk and deciles of predicted risk. The IPA combines discrimination and calibration in one summary value that ranges from 0 (poor) to 1 (perfect),^23^ and for this reason the IPA was used as the guiding measure for deriving the super learner’s coefficients. Net benefit is based on the weighted difference in the true versus false positive rate of a prediction model at a given decision threshold, i.e.,

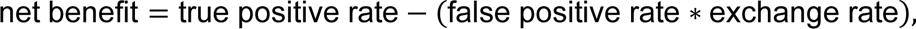

where the exchange rate is the odds of the threshold probability used for clinical decision making.^24^ Net benefit can be interpreted as the number of true-positive cases identified per 100 patients screened.^25^

Fairness was measured in terms of equal opportunity, equal odds, and demographic parity, with each metric based on equity in subgroups defined by race and sex.^26,27^ These metrics are computed by computing a score in each subgroup, then dividing the least favorable score by the most. For example, if patient in groups A, B, and C have 10%, 15%, and 20% chance, respectively, to be recommended to receive a treatment based on a prediction model, demographic parity would be computed using groups A and C and would be 10/20 = 50%. For each fairness metric, we considered a score < 80% to indicate lack of equity for at least one group. Decision thresholds of 2.5%, 5%, 7.5%, and 10% were used when assessing net benefit, fairness, sensitivity, specificity, and positive/negative predictive value.

Partial dependence was used to explain model predictions.^28^ Briefly, partial dependence allows for estimation of the expected prediction from a model when a single predictor or set of predictors take specific values. In the current analysis, we measured expected risk using the median value, and used the 25th and 75th percentile to quantify variability. Prior studies have shown partial dependence can produce biased explanations when relationships of predictors are not accounted for.^29^ We therefore focused explanations of partial dependence to consider sequences of diagnoses that frequently occur in clinical settings.

All analyses were performed using R version 4.2.3 (R Project for Statistical Computing [http://www.r-project.org]) with assistance from multiple R packages.^30–35^ All hypothesis tests were 2-sided, and P values less than 0.05 were considered statistically significant.

## RESULTS

We included 50,553 and 72,080 patients in the training and testing data, respectively (Table S2). Patients in the training versus testing data were slightly younger, with a mean age of 64.3 versus 65.6 years (Table 1). The median (25th, 75th percentile) number of days available as a look-back period was 815 (583, 1,099) for patients in the training data and 700 (419, 1,361) for patients in the testing data. A total of 1,820 and 721 incident MCI and dementia diagnoses, respectively, occurred over 209,259 person-years in the training cohort, with 2,066 and 729 incident MCI and dementia diagnoses, respectively, over 181,935 person-years in the testing cohort. The 3-year cumulative incidence (95% confidence interval [CI]) of MCI or dementia was 3.1% (3.0%, 3.3%) in the training data and 4.7% (4.5%, 4.9%) in the testing data (Figure S1).

**Table 1:** Patient characteristics before or on the index date, overall and stratified by cohort.

| **Characteristic\*** | **Overall**<br>**N = 122,633** | **Training data**<br>**N = 50,553** | **Testing data**<br>**N = 72,080** |
| --- | --- | --- | --- |
| Age, years | 65 (6) | 64 (6) | 66 (7) |
| Female | 57 | 57 | 57 |
| Race |  |  |  |
| White | 78 | 78 | 77 |
| Black | 19 | 18 | 19 |
| Other | 3.6 | 3.7 | 3.5 |
| Hispanic | 2.2 | 1.9 | 2.3 |
| Smoking habits |  |  |  |
| Never | 55 | 56 | 54 |
| Former | 34 | 34 | 35 |
| Current | 10 | 9.8 | 11 |
| Body mass index | 31 (7) | 31 (7) | 31 (7) |
| Systolic blood pressure | 130 (14) | 128 (14) | 131 (13) |
| Diastolic blood pressure | 76 (8) | 76 (8) | 76 (8) |
| Comorbidities<sup>†</sup> |  |  |  |
| Hypertension | 33 | 17 | 42 |
| Orthostatic hypotension | <0.1 | <0.1 | 0.1 |
| Atrial fibrillation | 4.4 | 2.0 | 6.0 |
| Cardiovascular disease | 0.6 | 0.1 | 0.9 |
| Stroke | 0.7 | 0.2 | 1.1 |
| Diabetes | 17 | 8.3 | 22 |
| Family history of AD | <0.1 | <0.1 | <0.1 |
| Depression | 0.9 | 0.3 | 1.4 |
| Head trauma | 1.9 | 0.9 | 2.5 |
| Insomnia | 2.8 | 1.2 | 3.9 |

\*Table values are mean (standard deviation) or percent
†Comorbidities indicated by diagnosis and presence in patient problem list prior to the index date.
Abbreviations: AD = Alzheimer's disease; EHR = electronic health records; and MCI = mild cognitive impairment

### Internal validation

Using both diagnosis codes and problem list indications to create predictors led to the highest IPA (4.3%) of cross-validated predictions (Table S3). The super learner that obtained this IPA combined predictions from the oblique RSF, gradient boosted trees, and standard Cox regression with coefficients of 0.867, 0.071, and 0.062, respectively. Thus, a ‘finalized’ super learner was created by replicating the workflow to fit these 3 models on the entire set of training data and then using the coefficients above to combine their predictions.

### External validation

The super learner obtained calibration intercept and slope (95% CI), respectively, of 0.000 (-0.008, 0.008) and 0.990 (0.821, 1.158) in the testing data (Figure 1). In subgroups based on sex and race, the super learner’s calibration intercept ranged from -0.002 (-0.018, 0.013) to 0.004 (-0.001, 0.009) and its calibration slope ranged from 0.954 (0.819, 1.089) to 1.165 (0.802, 1.529) (Figure S2). The super learner obtained a C-statistic (95% CI) of 0.811 (0.810, 0.812) in the testing data (Figure 2). Parity of the C-statistic for all learners exceeded 0.90 in subgroups based on race and sex.

**Figure 1:**
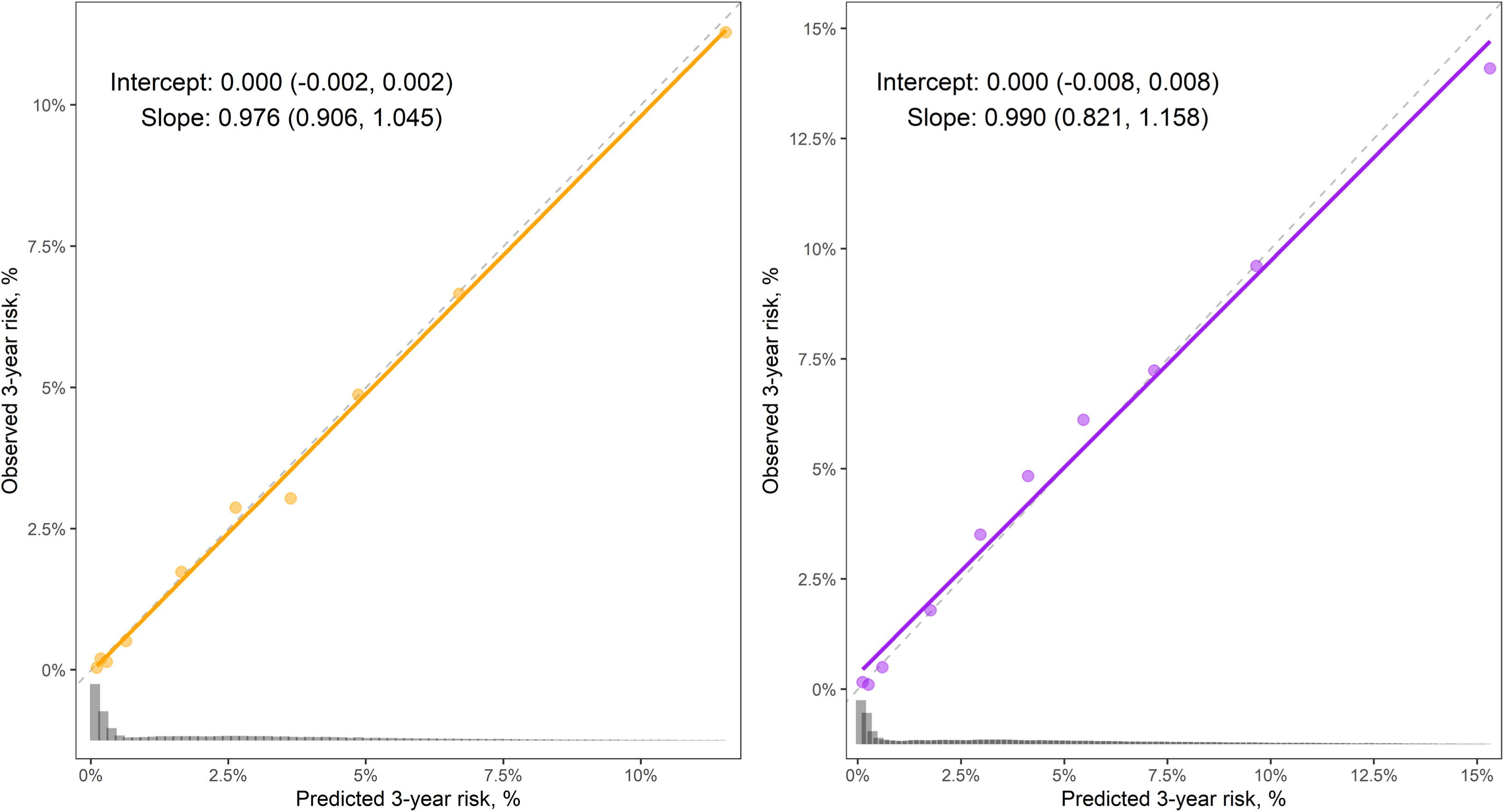
Calibration of the super learner in the training data (left panel) and testing data (right panel).

**Figure 2:**
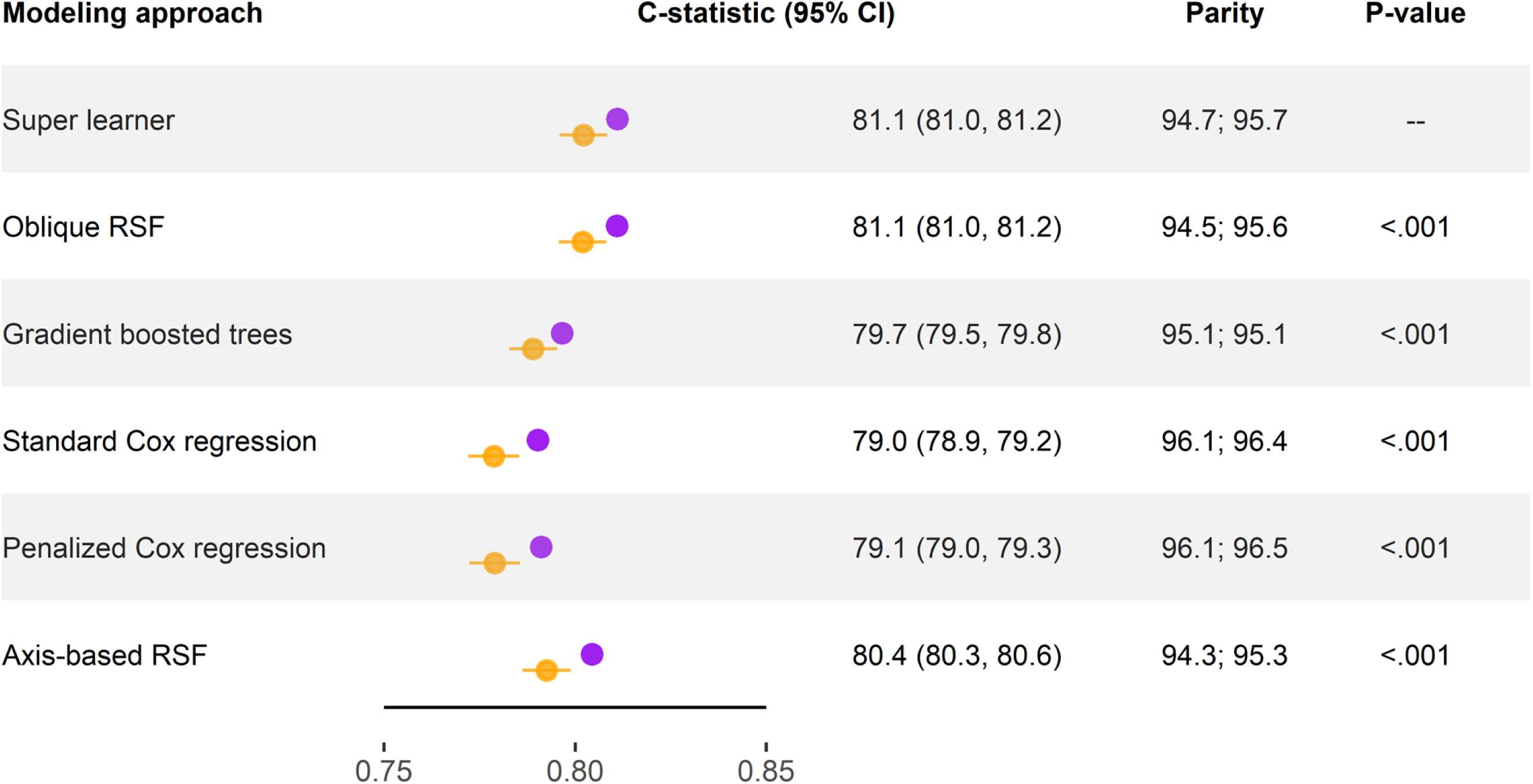
Concordance statistic and 95% confidence interval for several modeling approaches in the training data (orange points) and testing data (purple points). Parity is computed by dividing the concordance statistic for each protected subgroup within race and sex by the maximum concordance within that group. P-values are presented for a test of non-zero difference between the concordance statistic of the super learner in the testing data versus each of the other learners, separately.

Applying the super learner to predict risk in the testing data with a decision threshold set at 5% resulted in a net benefit of 18 true positive cases identified per 1000 patients (Figure 3). In subgroups based on race and sex, the net benefit using a decision threshold of 5% ranged from 15 for men to 20 for women (Figure S3).

**Figure 3:**
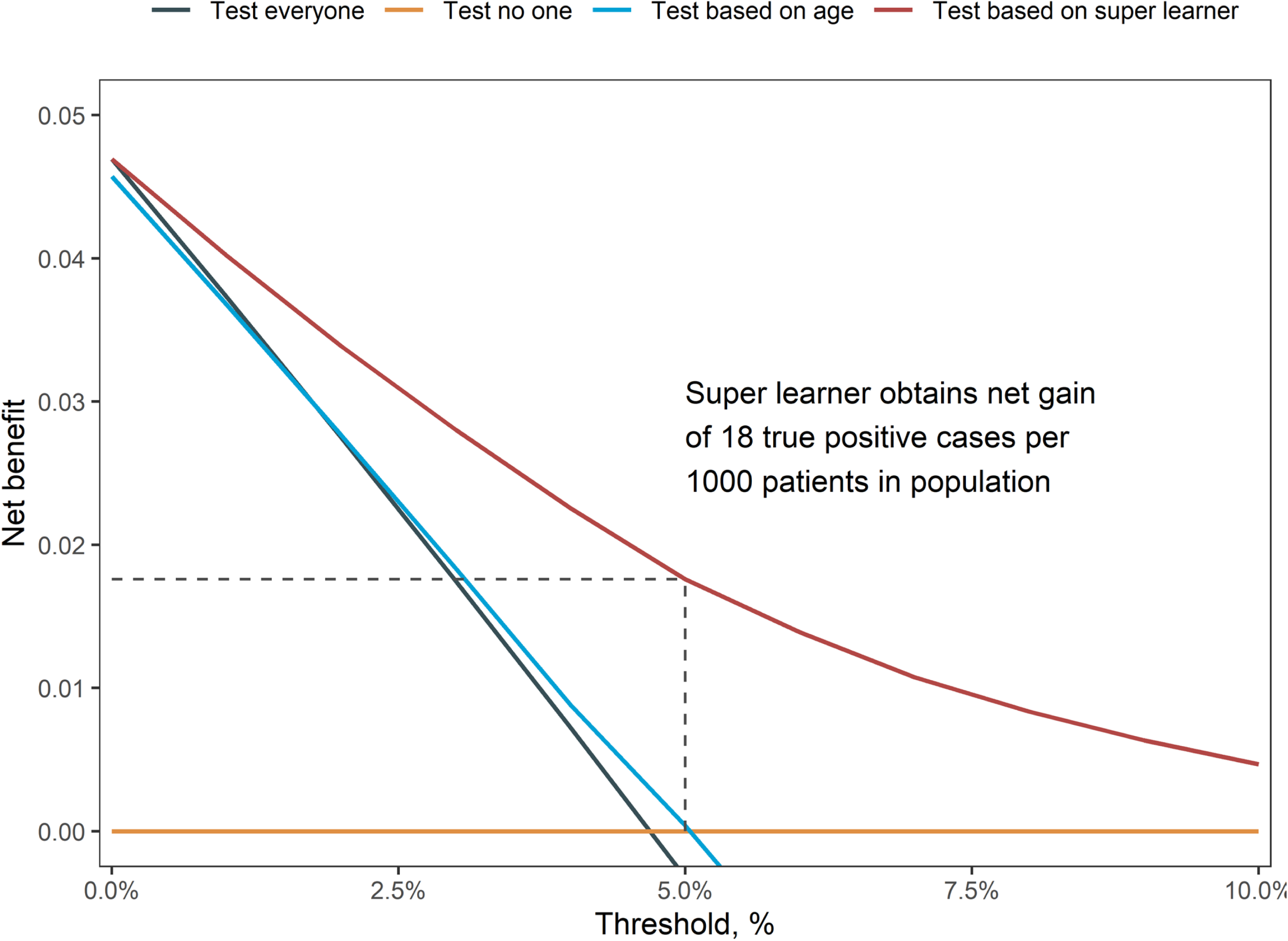
Decision curve analysis for several modeling approaches in the testing data.

### Fairness and testing characteristics

In both the training and testing data, decision-making using a risk threshold of 2.5% and 5% from the super learner maintained equal opportunity, equal odds, and demographic parity ratios of at least 73% and 83%, respectively, in subgroups based on race and sex, indicating acceptable fairness for the 5% threshold (Table 2).^26^ When the risk threshold was 7.5%, the super learner obtained a equal opportunity of 71% in internal data among subgroups based on sex, indicating slight bias. When the risk threshold was 10%, the super learner obtained equal opportunity of 65% in internal data among subgroups based on sex, indicating potential complications in fairness if this threshold were used in clinical settings. Using thresholds of 2.5, 5.0, 7.5, and 10, the overall percentage of patients who would be recommended to undergo cognitive screening was 56%, 33%, 18%, and 9.0%, respectively (Table S5).

**Table 2:** Fairness of the super learner’s predictions in patient subgroups based on Race and sex using multiple risk thresholds for clinical decision-making.

| Decision threshold, % | Training data |  |  | Testing data |  |  |
| --- | --- | --- | --- | --- | --- | --- |
|  | Equal opportunity* | Equal odds† | Demographic parity‡ | Equal opportunity | Equal odds | Demographic parity |
| Race |  |  |  |  |  |  |
| 2.5 | 99 | 82 | 81 | 98 | 73 | 83 |
| 5.0 | 99 | 95 | 84 | 97 | 93 | 90 |
| 7.5 | 86 | 86 | 88 | 89 | 89 | 99 |
| 10.0 | 77 | 77 | 91 | 78 | 78 | 92 |
| Sex |  |  |  |  |  |  |
| 2.5 | 95 | 93 | 93 | 95 | 92 | 95 |
| 5.0 | 83 | 83 | 83 | 83 | 83 | 85 |
| 7.5 | 71 | 71 | 71 | 74 | 74 | 74 |
| 10.0 | 65 | 65 | 65 | 73 | 73 | 66 |

\*Equal opportunity is satisfied when a model's predictions have the same true positive and false negative rates across protected groups.
†Equalized odds is satisfied when a model's predictions have the same false positive, true positive, false negative, and true negative rates across protected groups.
‡Demographic parity is satisfied when a model's predictions have the same predicted positive rate across groups.

### Variable importance and partial dependence

The three conditions that were ranked most important for prediction were hypertension, head trauma, and depression (Table 3). For male and female patients aged 65 years with hypertension diagnosis and problem list indication for 6 months, the median 3-year predicted risk (25th, 75th percentile) from the super learner was 5.8 (3.9, 6.7) and 6.1 (4.5, 7.0), respectively, while median 3-year predicted risk was 2.5 (0.35, 5.0) and 2.7 (0.33, 5.4) for male and female patients, respectively, aged 65 years with no hypertension. Increased frequency of hypertension diagnosis did not increase the median 3-year predicted risk from the super learner, whereas diagnosis and problem list indication of depression in 0, 10% and 20% of visits led to a median 3-year predicted risk of 2.9 (0.35, 5.2), 13 (11, 15), and 13 (12, 15), respectively, for males, and 3.2 (0.33, 5.7), 13 (12, 16), and 14 (13, 16), respectively, for females (Table S4). Partial dependence plots of the super learner’s 3-year predicted risk confirmed that common trajectories of diagnoses traditionally associated with dementia diagnosis conferred higher predicted risk for male and female patients (Figures S4-S6).

**Table 3:** Multi-variable adjusted 3-year predicted risk from the super learner for incident diagnosis of mild cognitive impairment or dementia based on the presence of different conditions for 6 months or 2 years.

| Type of indication | Aged 65 years |  | Aged 75 years |  |
| --- | --- | --- | --- | --- |
|  | Female | Male | Female | Male |
| Hypertension, variable importance = 10 |  |  |  |  |
| None | 2.7<br>(0.33, 5.4) | 2.5<br>(0.35, 5.0) | 5.7<br>(1.9, 8.4) | 5.5<br>(1.9, 8.0) |
| Diagnosis and problem list for 0.5 years | 4.9<br>(4.1, 6.3) | 4.4<br>(3.7, 5.7) | 9.3<br>(8.5, 11) | 9.2<br>(7.8, 11) |
| Diagnosis and problem list for 2 years | 6.1<br>(4.5, 7.0) | 5.8<br>(3.9, 6.7) | 12<br>(9.9, 13) | 12<br>(8.9, 13) |
| Head trauma, variable importance = 3.5 |  |  |  |  |
| None | 3.2<br>(0.33, 5.5) | 2.9<br>(0.35, 5.1) | 6.8<br>(1.9, 10) | 6.1<br>(1.9, 9.7) |
| Diagnosis and problem list for 0.5 years | 7.2<br>(6.3, 9.1) | 6.9<br>(6.1, 8.7) | 11<br>(9.3, 14) | 11<br>(9.2, 14) |
| Diagnosis and problem list for 2 years | 8.0<br>(7.2, 9.9) | 7.7<br>(6.9, 9.7) | 12<br>(11, 15) | 12<br>(10, 15) |
| Depression, variable importance = 0.88 |  |  |  |  |
| None | 3.2<br>(0.33, 5.7) | 2.9<br>(0.35, 5.2) | 6.8<br>(1.9, 11) | 6.1<br>(1.9, 9.8) |
| Diagnosis and problem list for 0.5 years | 5.0<br>(3.9, 7.2) | 4.7<br>(3.9, 6.7) | 9.0<br>(6.2, 13) | 8.5<br>(6.4, 12) |
| Diagnosis and problem list for 2 years | 5.5<br>(4.4, 7.7) | 5.3<br>(4.3, 7.2) | 9.5<br>(6.9, 13) | 9.0<br>(7.2, 12) |
| Stroke, variable importance = 0.36 |  |  |  |  |
| None | 3.2<br>(0.33, 5.7) | 2.9<br>(0.35, 5.2) | 6.8<br>(1.9, 11) | 6.1<br>(1.9, 9.9) |
| Diagnosis and problem list for 0.5 years | 5.8<br>(3.9, 8.2) | 5.7<br>(3.9, 7.9) | 10<br>(7.0, 14) | 10<br>(7.5, 14) |
|  | Female | Male | Female | Male |
| Diagnosis and problem list for 2 years | 6.2<br>(4.0, 8.7) | 6.1<br>(4.1, 8.4) | 11<br>(7.4, 15) | 11<br>(8.3, 14) |
| Orthostatic hypotension, variable importance = 0.15 |  |  |  |  |
| None | 3.2<br>(0.33, 5.7) | 2.9<br>(0.35, 5.3) | 6.8<br>(1.9, 11) | 6.1<br>(1.9, 10) |
| Diagnosis and problem list for 0.5 years | 5.1<br>(3.2, 7.9) | 4.8<br>(3.1, 7.4) | 10<br>(6.1, 13) | 9.3<br>(5.8, 13) |
| Diagnosis and problem list for 2 years | 5.3<br>(3.5, 8.2) | 5.0<br>(3.5, 7.7) | 10<br>(6.5, 14) | 9.6<br>(6.3, 13) |
| Cardiovascular disease, variable importance = 0.13 |  |  |  |  |
| None | 3.2<br>(0.33, 5.7) | 2.9<br>(0.35, 5.2) | 6.8<br>(1.9, 11) | 6.1<br>(1.9, 9.9) |
| Diagnosis and problem list for 0.5 years | 6.5<br>(2.6, 9.5) | 6.0<br>(2.4, 8.8) | 11<br>(5.5, 15) | 10<br>(5.8, 14) |
| Diagnosis and problem list for 2 years | 6.7<br>(2.8, 9.9) | 6.2<br>(2.4, 9.2) | 11<br>(5.9, 16) | 11<br>(6.2, 15) |
| Family history of AD, variable importance = 0.02 |  |  |  |  |
| None | 3.2<br>(0.33, 5.7) | 2.9<br>(0.35, 5.2) | 6.8<br>(1.9, 11) | 6.1<br>(1.9, 10) |
| Diagnosis and problem list for 0.5 years | 7.4<br>(5.6, 11) | 6.8<br>(5.2, 10) | 12<br>(8.6, 17) | 11<br>(8.1, 15) |
| Diagnosis and problem list for 2 years | 7.8<br>(5.9, 12) | 7.2<br>(5.6, 11) | 13<br>(8.9, 17) | 12<br>(8.5, 16) |

## DISCUSSION

We leveraged structured EHR data to develop a ML prediction model for the 3-year risk of an incident diagnosis for MCI or dementia for persons actively seen in a healthcare system. In prospective validation, the model obtained a C-statistic of 0.811 (0.810, 0.812) with adequate calibration overall and in subgroups based on race and sex. Recommending screening to patients with 3-year predicted risk of 5% or higher provided a net benefit of 18 true positive cases identified per 1000 patients while maintaining equal opportunity, equal odds, and demographic parity values over 80% in subgroups based on race and sex. These results provide evidence that ML-based algorithms within the EHR can produce recommendations for earlier cognitive assessment of patients that have high probability of being diagnosed with cognitive impairment within three years. With emerging clinical trial evidence for both behavioral and pharmacologic interventions to slow the progression of various etiologies of MCI and dementia, development of better tools for identification of adults at high risk of these conditions increases the opportunity for patient participation in preventive interventions.

Several other studies have demonstrated the potential for ML to improve risk prediction for cognitive impairment.^36^ While the super learner model developed in this work generally demonstrates comparable predictive performance, in terms of discrimination, there are several relevant pros and cons to consider. The performance of our model comes with the benefit of simplicity, as many ML models have leveraged hundreds of predictors from structured and unstructured EHR data, making the algorithms more complex and less scalable.^6,7^ As far as we are aware, no existing algorithms have systematically evaluated algorithmic fairness with respect to sex and race, with many algorithms failing to even report the racial/ethnic composition of the samples on which they were developed.^36^ The assessment of fairness and equity has high importance given historical disparities in dementia diagnosis and care.^37–39^ The primary limitation of our work is the lack of a reference standard for model development. While the model demonstrates reasonable performance, it is relative to an outcome (clinical diagnoses of MCI or dementia) that is known to under-report the true burden of disease. With that said, this limitation may be less relevant for applications like prioritizing cognitive screening within medical practice, versus other applications that have tried to identify prevalent cognitive impairment that had not been clinically documented.^40^

A scalable model to predict risk for cognitive impairment could be useful in a variety of health care settings, particularly primary care, recognizing the myriad of barriers to conducting additional screening in that setting.^41^ The primary goal of our work is to identify at-risk individuals that may or may not have indicated a subjective cognitive complaint. The finalized model from the current analysis is currently integrated within the AH-WFB EHR, with 3-year predicted risk probabilities generated on a monthly basis. The score is being prospectively validated as part of a research study funded by the Alzheimer’s Association evaluating a pathway in primary care that includes referral for telephone cognitive testing and the assessment of blood-based biomarkers for Alzheimer’s disease. The pathway after receiving a predicted score from the current model may include subsequent referrals to several possible interventions depending on the pathology underlying any identified cognitive impairment.

The current study has several strengths. We leveraged a large, contemporary cohort and performed prospective, temporal validation of a prediction model developed using the “super learner” technique. We systematically explored the prognostic value of multiple initial predictor sets (diagnoses, problem lists, or both) using internal validation. Our final model obtained adequate discrimination and calibration while only leveraging diagnosis codes observed prior to an index date to generate predictions. We assessed the fairness of our final model in subgroups based on race and sex. The current study also has several known limitations. The time period used to define our model’s training data included the onset of the COVID-19 pandemic, which may have impacted diagnoses of MCI and dementia.^42^ The median look-back period to derive patient predictors in the current study was approximately 2 years, and this may not be a sufficient amount of time to adequately characterize medical history. The primary outcome was based in part on MCI diagnoses in the primary care setting, which is a somewhat unstable condition due to multi-dimensional frictions such as transient medical conditions that impact cognitive function and barriers associated with fully evaluating individuals’ cognitive performance.^43^

In summary, we developed an effective and fair ML prediction model for incident diagnoses of MCI or dementia diagnosis using routine structured EHR data. While the ML model requires prospective testing in combination with cognitive screening, the hope is that it provides an efficient mechanism to identify individuals at high risk for cognitive impairment, potentially facilitating earlier screening and intervention.

## Data Availability

The data used in this study were obtained from patient medical records and are not publicly available due to privacy and confidentiality restrictions.

## CONSENT STATEMENT

This research was approved by the Wake Forest University Health Sciences Institutional Review Board under a waiver of informed consent.

## REFERENCES

1. Alzheimer’s Disease Facts and Figures. Alzheimer’s Disease and Dementia. Accessed March 4, 2024. https://www.alz.org/alzheimers-dementia/facts-figures

2. Owens DK, Davidson KW, Krist AH, et al. Screening for cognitive impairment in older adults: US preventive services task force recommendation statement. Jama. 2020;323(8):757–763.

3. Van Dyck CH, Swanson CJ, Aisen P, et al. Lecanemab in early alzheimer’s disease. New England Journal of Medicine. 2023;388(1):9–21.

4. Sims JR, Zimmer JA, Evans CD, et al. Donanemab in early symptomatic alzheimer disease: The TRAILBLAZER-ALZ 2 randomized clinical trial. Jama. 2023;330(6):512–527.

5. Williamson JD, Pajewski NM, Auchus AP, et al. Effect of intensive vs standard blood pressure control on probable dementia: A randomized clinical trial. Jama. 2019;321(6):553–561.

6. Fouladvand S, Noshad M, Periyakoil V, Chen JH. Machine learning prediction of mild cognitive impairment and its progression to alzheimer’s disease. Health Science Reports. 2023;6(10):e1438.

7. You J, Zhang YR, Wang HF, et al. Development of a novel dementia risk prediction model in the general population: A large, longitudinal, population-based machine-learning study. EClinicalMedicine. 2022;53. Accessed March 5, 2024. https://www.thelancet.com/journals/eclinm/article/PIIS2589-5370(22)00395-9/fulltext

8. Orkaby AR, Callahan KE, Driver JA, Hudson K, Clegg AJ, Pajewski NM. New horizons in frailty identification via electronic frailty indices: Early implementation lessons from experiences in england and the united states. Age and ageing. 2024;53(2):afae025.

9. Simon N, Friedman J, Hastie T, Tibshirani R. Regularization paths for cox’s proportional hazards model via coordinate descent. Journal of Statistical Software. 2011;39(5):1–13. http://www.jstatsoft.org/v39/i05/

10. Jaeger BC, Long DL, Long DM, et al. Oblique random survival forests. The Annals of Applied Statistics. 2019;13(3):1847–1883. doi:10.1214/19-AOAS1261

11. Jaeger BC, Welden S, Lenoir K, et al. Accelerated and interpretable oblique random survival forests. Journal of Computational and Graphical Statistics. Published online 2023:1–16.

12. Chen T, He T, Benesty M, et al. Xgboost: Extreme Gradient Boosting.; 2022. https://CRAN.R-project.org/package=xgboost

13. Van der Laan MJ, Polley EC, Hubbard AE. Super learner. Statistical applications in genetics and molecular biology. 2007;6(1).

14. Polley EC, Van der Laan MJ. Super learner in prediction. Published online 2010.

15. Degenhardt F, Seifert S, Szymczak S. Evaluation of variable selection methods for random forests and omics data sets. Briefings in bioinformatics. 2019;20(2):492–503.

16. Speiser JL, Miller ME, Tooze J, Ip E. A comparison of random forest variable selection methods for classification prediction modeling. Expert systems with applications. 2019;134:93–101.

17. Breiman L. Random forests. Machine learning. 2001;45:5–32.

18. Menze BH, Kelm BM, Splitthoff DN, Koethe U, Hamprecht FA. On oblique random forests. In: Machine Learning and Knowledge Discovery in Databases: European Conference, ECML PKDD 2011, Athens, Greece, September 5-9, 2011, Proceedings, Part II 22. Springer; 2011:453–469.

19. Hastie T, Tibshirani R, Friedman JH, Friedman JH. The wrong and right way to do cross-validation. In: The Elements of Statistical Learning: Data Mining, Inference, and Prediction. Vol 2. Springer; 2009:245–247.

20. Steyerberg EW, Vickers AJ, Cook NR, et al. Assessing the performance of prediction models: A framework for some traditional and novel measures. *Epidemiology (Cambridge*, Mass). 2010;21(1):128.

21. Pessach D, Shmueli E. A review on fairness in machine learning. ACM Computing Surveys (CSUR). 2022;55(3):1–44.

22. Blanche P, Dartigues JF, Jacqmin-Gadda H. Estimating and comparing time-dependent areas under receiver operating characteristic curves for censored event times with competing risks. Statistics in medicine. 2013;32(30):5381–5397.

23. Kattan MW, Gerds TA. The index of prediction accuracy: An intuitive measure useful for evaluating risk prediction models. Diagnostic and prognostic research. 2018;2:1–7.

24. Vickers AJ, Cronin AM, Elkin EB, Gonen M. Extensions to decision curve analysis, a novel method for evaluating diagnostic tests, prediction models and molecular markers. BMC medical informatics and decision making. 2008;8:1–17.

25. Vickers AJ, Calster B van, Steyerberg EW. A simple, step-by-step guide to interpreting decision curve analysis. Diagnostic and prognostic research. 2019;3:1–8.

26. Allen C, Ahmad MA, Eckert C, Hu J, Kumar V, Teredesai A. fairMLHealth: Tools and tutorials for fairness evaluation in healthcare machine learning. GitHub repository. Published online 2020.

27. Evans C, Johnson E, Lin J. Assessing algorithmic bias and fairness in clinical prediction models for preventive services. A health equity methods project for the US preventive services task force. 2023.

28. Friedman JH. Greedy function approximation: A gradient boosting machine. The Annals of Statistics. 2001;29(5):1189–1232. Accessed May 16, 2024. http://www.jstor.org/stable/2699986

29. Molnar C, König G, Herbinger J, et al. General pitfalls of model-agnostic interpretation methods for machine learning models. In: Holzinger A, Goebel R, Fong R, Moon T, Müller KR, Samek W, eds. AI - Beyond Explainable AI: International Workshop, Held in Conjunction with ICML 2020, July 18, 2020, Vienna, Austria, Revised and Extended Papers. Springer International Publishing; 2022:39–68. doi:10.1007/978-3-031-04083-2_4

30. R Core Team. R: A Language and Environment for Statistical Computing. R Foundation for Statistical Computing; 2020. https://www.R-project.org/

31. Jaeger B. Table.glue: Make and Apply Customized Rounding Specifications for Tables. https://github.com/bcjaeger/table.glue

32. Wickham H, Averick M, Bryan J, et al. Welcome to the tidyverse. Journal of Open Source Software. 2019;4(43):1686. doi:10.21105/joss.01686

33. Therneau TM. A Package for Survival Analysis in R.; 2021. https://CRAN.R-project.org/package=survival

34. Landau WM. The targets R package: A dynamic make-like function-oriented pipeline toolkit for reproducibility and high-performance computing. Journal of Open Source Software. 2021;6(57):2959. 10.21105/joss.02959

35. Jaeger BC, Welden S, Lenoir K, Pajewski NM. Aorsf: An R package for supervised learning using the oblique random survival forest. Journal of Open Source Software. 2022;7(77):4705.

36. Taylor B, Barboi C, Boustani M. Passive digital markers for alzheimer’s disease and other related dementias: A systematic evidence review. Journal of the American Geriatrics Society. 2023;71(9):2966–2974.

37. Gianattasio KZ, Prather C, Glymour MM, Ciarleglio A, Power MC. Racial disparities and temporal trends in dementia misdiagnosis risk in the united states. Alzheimer’s & Dementia: Translational Research & Clinical Interventions. 2019;5:891–898.

38. Lin PJ, Daly AT, Olchanski N, et al. Dementia diagnosis disparities by race and ethnicity. Medical care. 2021;59(8):679–686.

39. Kawas CH, Corrada MM, Whitmer RA. Diversity and disparities in dementia diagnosis and care: A challenge for all of us. JAMA neurology. 2021;78(6):650–652.

40. Barnes DE, Zhou J, Walker RL, et al. Development and validation of eRADAR: A tool using EHR data to detect unrecognized dementia. Journal of the American Geriatrics Society. 2020;68(1):103–111.

41. Porter J, Boyd C, Skandari MR, Laiteerapong N. Revisiting the time needed to provide adult primary care. Journal of general internal medicine. 2023;38(1):147–155.

42. Hoang M, Jurado P, Abzhandadze T, et al. Effects of the COVID-19 pandemic on the number of new dementia diagnoses and the quality of dementia diagnostics and treatment. The Journal of Prevention of Alzheimer’s Disease. Published online 2024:1–9.

43. Sabbagh MN, Boada M, Borson S, et al. Early Detection of Mild Cognitive Impairment (MCI) in Primary Care. The Journal of Prevention of Alzheimer’s Disease. 2020;7(3):165–170. doi:10.14283/jpad.2020.21

