## Supplemental tables and figures for "Design and Validation of a Pragmatic, Scalable Prioritization Tool for Cognitive Screening using the Electronic Health Record"

### SUPPLEMENT

Table S1: International Classification of Disease codes used to identify outcomes and comorbidities.

| **Condition** | **ICD-10 diagnostic codes** |
| --- | --- |
| Dementia | F01.A0, F01.A11, F01.A18, F01.A3, F01.A4, F01.B0, F01.B11, F01.B18, F01.B2, F01.B3, F01.B4, F01.C0, F01.C11, F01.C3, F02.A0, F02.A11, F02.A18, F02.A4, F02.B0, F02.B11, F02.B18, F02.B2, F02.B3, F02.B4, F02.C4, F03.A0, F03.A11, F03.A18, F03.A2, F03.A3, F03.A4, F03.B0, F03.B11, F03.B18, F03.B2, F03.B3, F03.B4, F03.C0, F03.C11, F03.C18, F03.C2, F03.C3, F03.C4, F05, F07.81, F10.97, G20, A81.0, F01.5, F02.8, F03.9, F06.8, F10.27, F19.97, G10, G11.9, G23, G30, G31.1, G31.2, G31.83, G31.89, G31.9, G90.3 |
| Mild cognitive impairment | G31.84, R41.3 |
| Atrial fibrillation | I48, I48.19, I48.20, I48.21, I48.91 |
| Cardiovascular disease | I67, I67.4, I67.8, I67.858, I67.89, I67.9, I69, I69.8, I69.80, I69.81, I69.810, I69.811, I69.812, I69.813, I69.814, I69.815, I69.818, I69.819, I69.820, I69.821, I69.822, I69.823, I69.828, I69.89, I69.890, I69.891, I69.892, I69.893, I69.898, I69.9, I69.90, I69.91, I69.910, I69.911, I69.912, I69.913, I69.914, I69.915, I69.918, I69.919, I69.92, I69.920, I69.921, I69.922, I69.923, I69.928, I69.99, I69.990, I69.991, I69.992, I69.993, I69.998, Z82.49 |
| Depression | F32.0, F32.1, F32.2, F32.3, F32.4, F32.5, F32.8, F32.89, F32.9 |
| Diabetes | E10, E11, E12, E13, E14 |
| Family history of Dementia | Z81.8, Z86.59 |
| Head trauma | F09, G44.309, R26.89, R40.20, R42, R45.89, R46.89, R53.83, S09.8XXA, S09.8XXD, S09.90XA, S09.90XS, S09.93XA, S09.93XD, S09.93XS |
| Hypertension | G93.2, H40.05, I10, I97.3, IMO0001, R03.0 |
| Insomnia | F13.982, F51.01, F51.02, F51.03, F51.04, F51.05, F51.09, G47.00, G47.01, G47.09, Z82.0 |
| Orthostatic hypotension | G90.3, I95.1 |
| Stroke | I67.2, I69.30, Z82.3, Z86.73 |

Table S2: Patients included in the current analysis.

| **Inclusion criteria** | **Number of patients** |
| --- | --- |
| Patients in EHR database | 629,566 |
| With >= 1 visit during index period^*^ | 305,823 |
| With >= 1 valid index date^†^ | 128,213 |
| With >= 1 visits after index date | 128,190 |
| ^*^The index period was defined as 01/01/2019 through 07/01/2021 for the training cohort and 07/02/2021 through 07/01/2022 for the testing cohort. | |
| ^†^The first office visit that occurred during the index period | |
| Abbreviations: AD = Alzheimer's disease; EHR = electronic health records; and MCI = mild cognitive impairment | |

Table S3: Indices of prediction accuracy and coefficients selected for inclusion in the super learner.

| **Model** | **Problem list and diagnoses** | | **Diagnoses only** | | **Problem list only** | |
| --- | --- | --- | --- | --- | --- | --- |
|  | **IPA** | **Coefficient** | **IPA** | **Coefficient** | **IPA** | **Coefficient** |
| Axis-based RSF | 3.7 | 0.00000000 | 3.4 | 0.00000000 | 1.9 | 0.21342538 |
| Penalized Cox regression | 3.1 | 0.00000000 | 3.0 | 0.00000000 | 2.0 | 0.04098228 |
| Standard Cox regression | 3.0 | 0.06201291 | 3.0 | 0.08734518 | 1.9 | 0.36604187 |
| Gradient boosted trees | 3.4 | 0.07091069 | 3.4 | 0.11601732 | 1.6 | 0.00000000 |
| Oblique RSF | 4.3 | 0.86707640 | 4.1 | 0.79663749 | 2.0 | 0.37955047 |
| Super learner | 4.3 |  | 4.1 |  | 2.2 |  |
| Abbreviations: IPA = index of prediction accuracy | | | | | | |

Table S4: Multi-variable adjusted 3-year predicted risk from the super learner for incident mild cognitive impairment or dementia based on the relative frequency of different indications.

| **Type of indication** | **Aged 65 years** | | **Aged 75 years** | |
| --- | --- | --- | --- | --- |
|  | **Female** | **Male** | **Female** | **Male** |
| Hypertension, variable importance = 10 | | | | |
| None | 5.5 (0.33, 8.4) | 5.0 (0.35, 7.7) | 9.7 (1.9, 13) | 9.3 (1.9, 12) |
| Diagnosis and problem list in 10% of visits | 4.1 (3.1, 5.7) | 3.8 (2.8, 5.3) | 7.8 (6.1, 9.9) | 6.9 (5.5, 8.9) |
| Diagnosis and problem list in 20% of visits | 4.2 (3.1, 5.8) | 3.8 (2.9, 5.3) | 7.8 (6.1, 9.8) | 6.9 (5.5, 8.9) |
| Depression, variable importance = 0.88 | | | | |
| None | 3.2 (0.33, 5.7) | 2.9 (0.35, 5.2) | 6.8 (1.9, 10) | 6.1 (1.9, 9.8) |
| Diagnosis and problem list in 10% of visits | 13 (12, 16) | 13 (11, 15) | 18 (16, 20) | 17 (15, 20) |
| Diagnosis and problem list in 20% of visits | 14 (13, 16) | 13 (12, 15) | 18 (16, 20) | 17 (16, 20) |

Table S5: Testing characteristics of the super learner with different thresholds for decision making.

| **Decision threshold, %** | **Test characteristic, %** | | | | |
| --- | --- | --- | --- | --- | --- |
|  | **Test positive rate** | **Sensitivity** | **Specificity** | **Positive predictive value** | **Negative predictive value** |
| Overall (N = 72,080) | | | | | |
| 2.5 | 56 | 93 | 46 | 7.8 | 99 |
| 5.0 | 33 | 71 | 69 | 10 | 98 |
| 7.5 | 18 | 47 | 84 | 12 | 97 |
| 10 | 9.0 | 29 | 92 | 15 | 96 |
| White patients (N = 54,488) | | | | | |
| 2.5 | 53 | 92 | 48 | 7.9 | 99 |
| 5.0 | 32 | 72 | 70 | 10 | 98 |
| 7.5 | 17 | 47 | 84 | 12 | 97 |
| 10 | 8.8 | 30 | 92 | 16 | 96 |
| Black patients (N = 13,484) | | | | | |
| 2.5 | 64 | 94 | 38 | 7.6 | 99 |
| 5.0 | 36 | 69 | 66 | 9.9 | 97 |
| 7.5 | 18 | 43 | 84 | 12 | 96 |
| 10 | 8.2 | 22 | 93 | 14 | 96 |
| Female (N = 40,732) | | | | | |
| 2.5 | 55 | 96 | 47 | 8.3 | 100 |
| 5.0 | 36 | 78 | 66 | 10 | 98 |
| 7.5 | 21 | 52 | 81 | 12 | 97 |
| 10 | 11 | 34 | 90 | 15 | 96 |
| Male (N = 31,348) | | | | | |
| 2.5 | 57 | 89 | 45 | 7.2 | 99 |
| 5.0 | 29 | 62 | 72 | 9.7 | 98 |
| 7.5 | 14 | 40 | 87 | 13 | 97 |
| 10 | 6.8 | 23 | 94 | 16 | 96 |

Figure S1: Cumulative incidence of dementia diagnosis in the training and testing data.

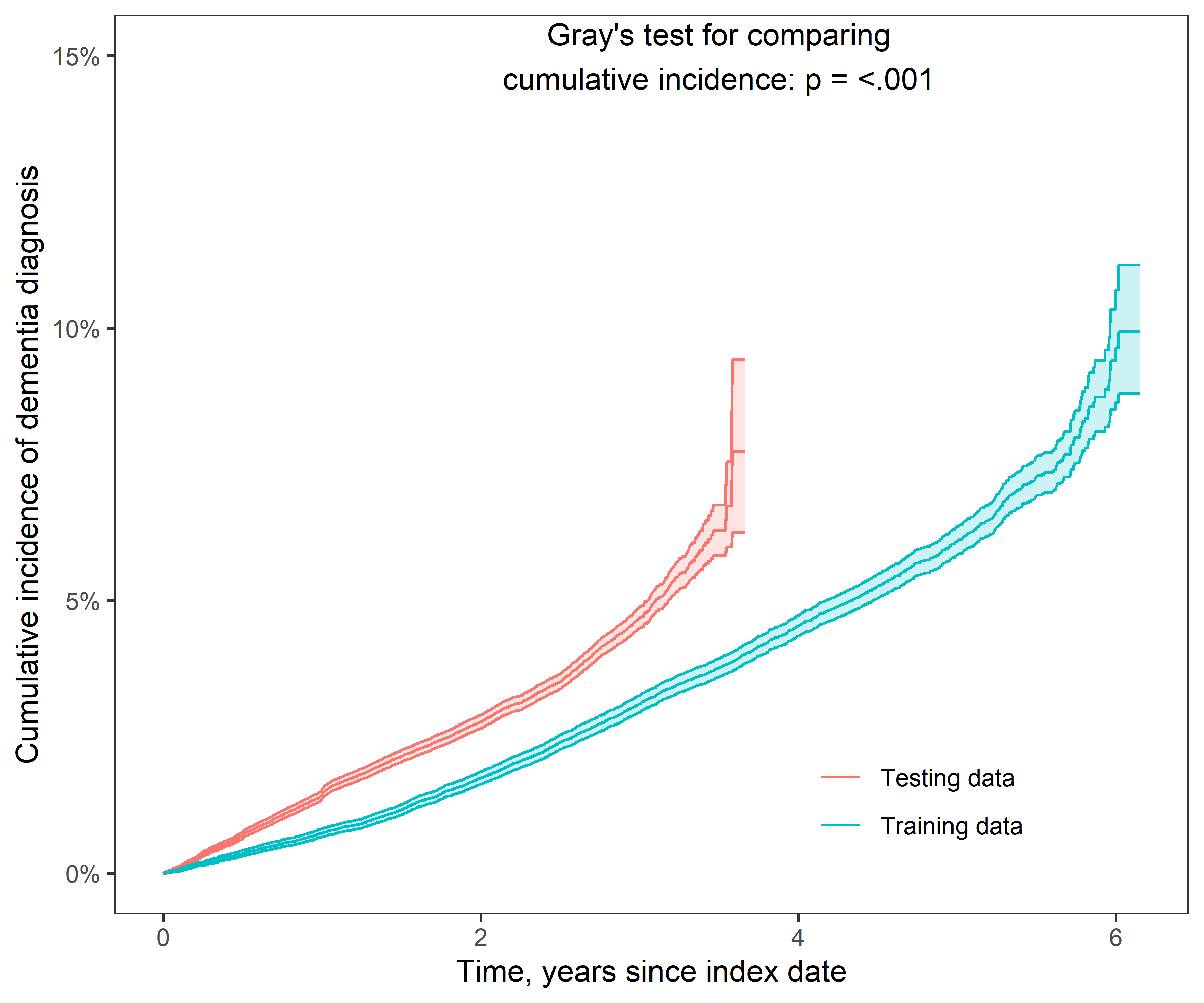

Figure S2: Calibration of the super learner in subsets of the testing data based on sex and race.

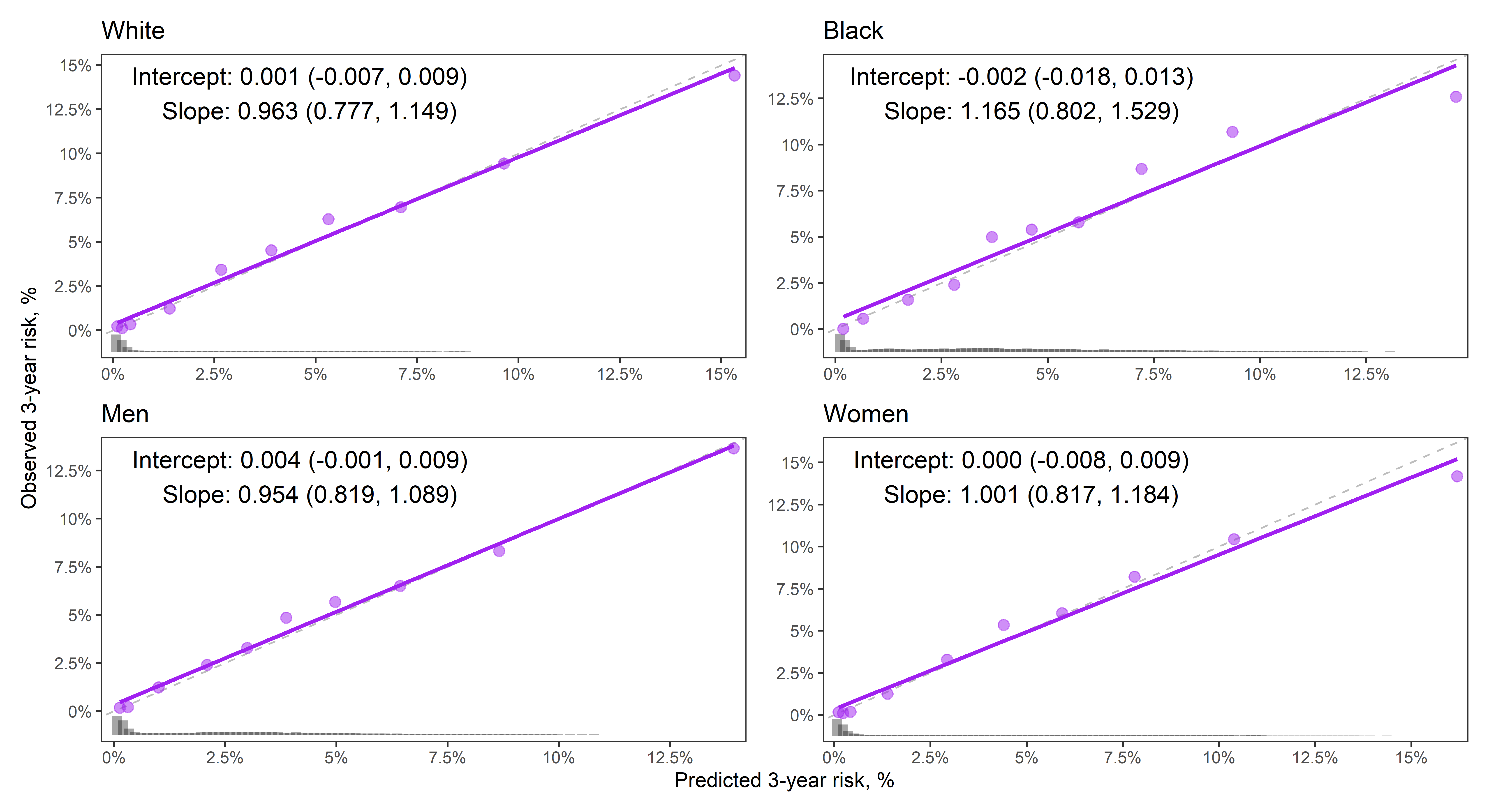

Figure S3: Decision curve analysis for several modeling approaches in the testing data among subgroups based on sex and race.

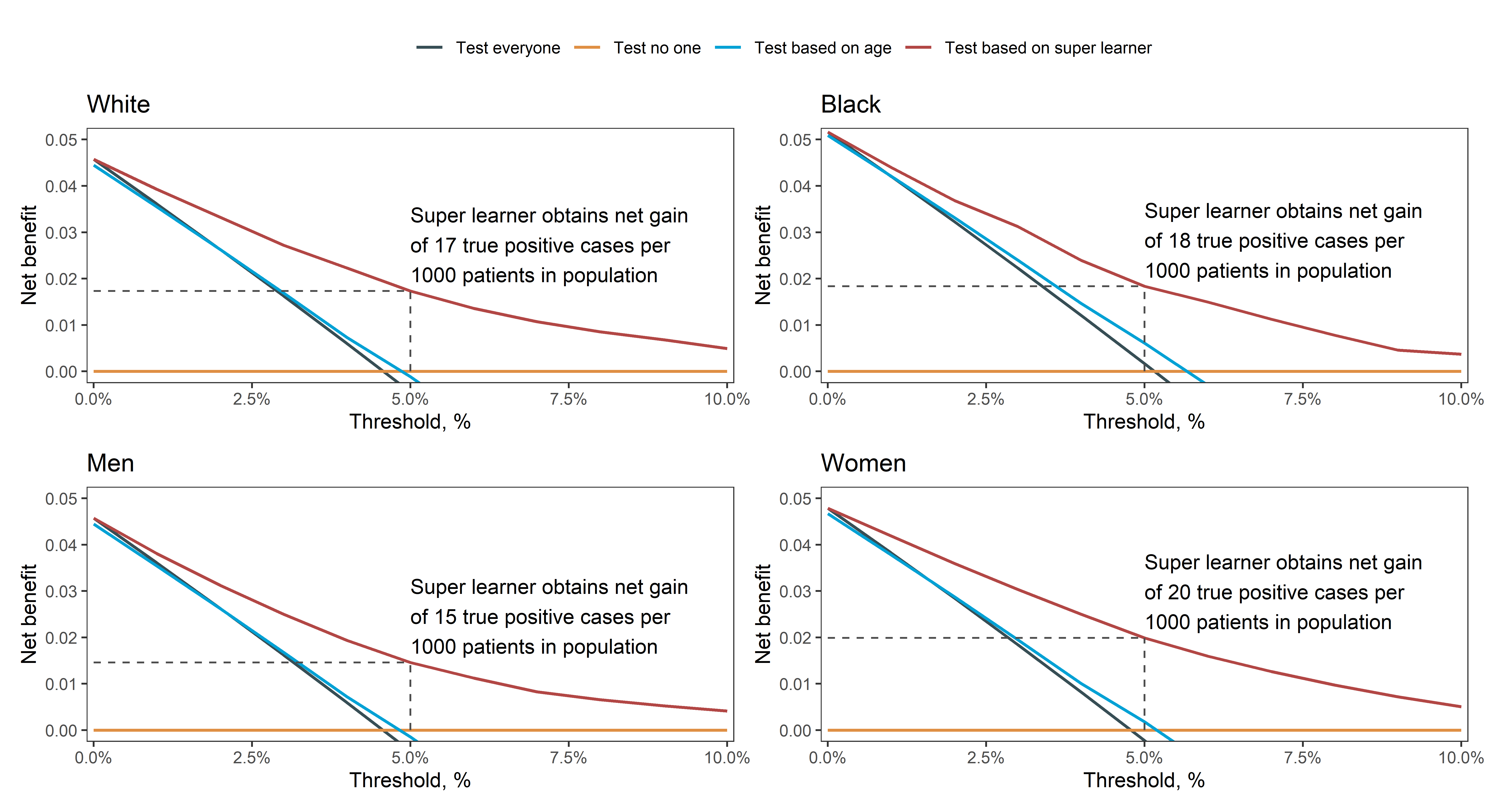

Figure S4: Multivariable adjusted 3-year predicted risk (25th, 75th percentile) for male and female patients aged 65 and 75 years. Prior to the index date, patients were diagnosed with hypertension (first row), hypertension and stroke (second row), or hypertension, stroke, and depression (third row).

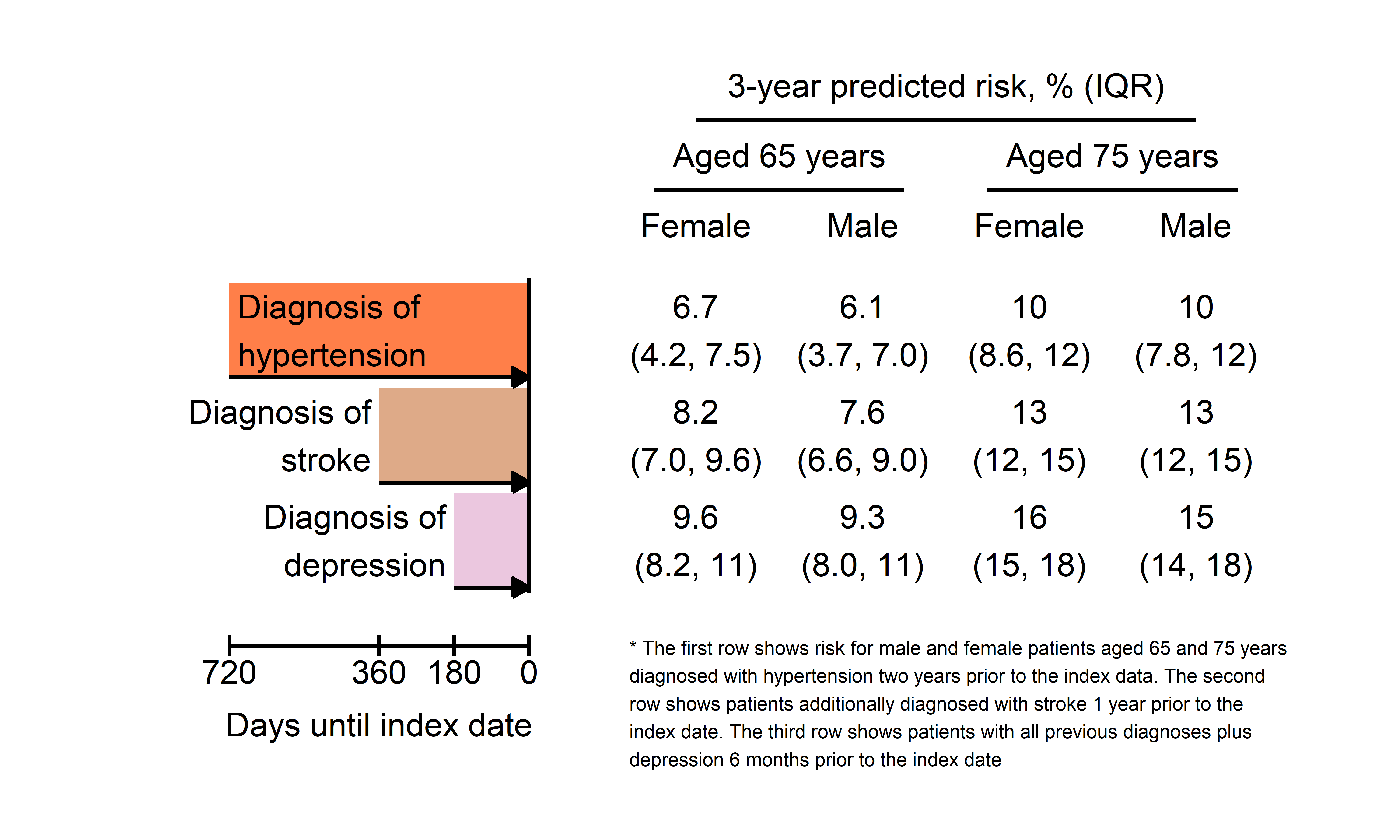

Figure S5: Multivariable adjusted 3-year predicted risk (25th, 75th percentile) for male and female patients aged 65 and 75 years. Prior to the index date, patients were diagnosed with head trauma (first row) or head trauma and insomnia (second row).

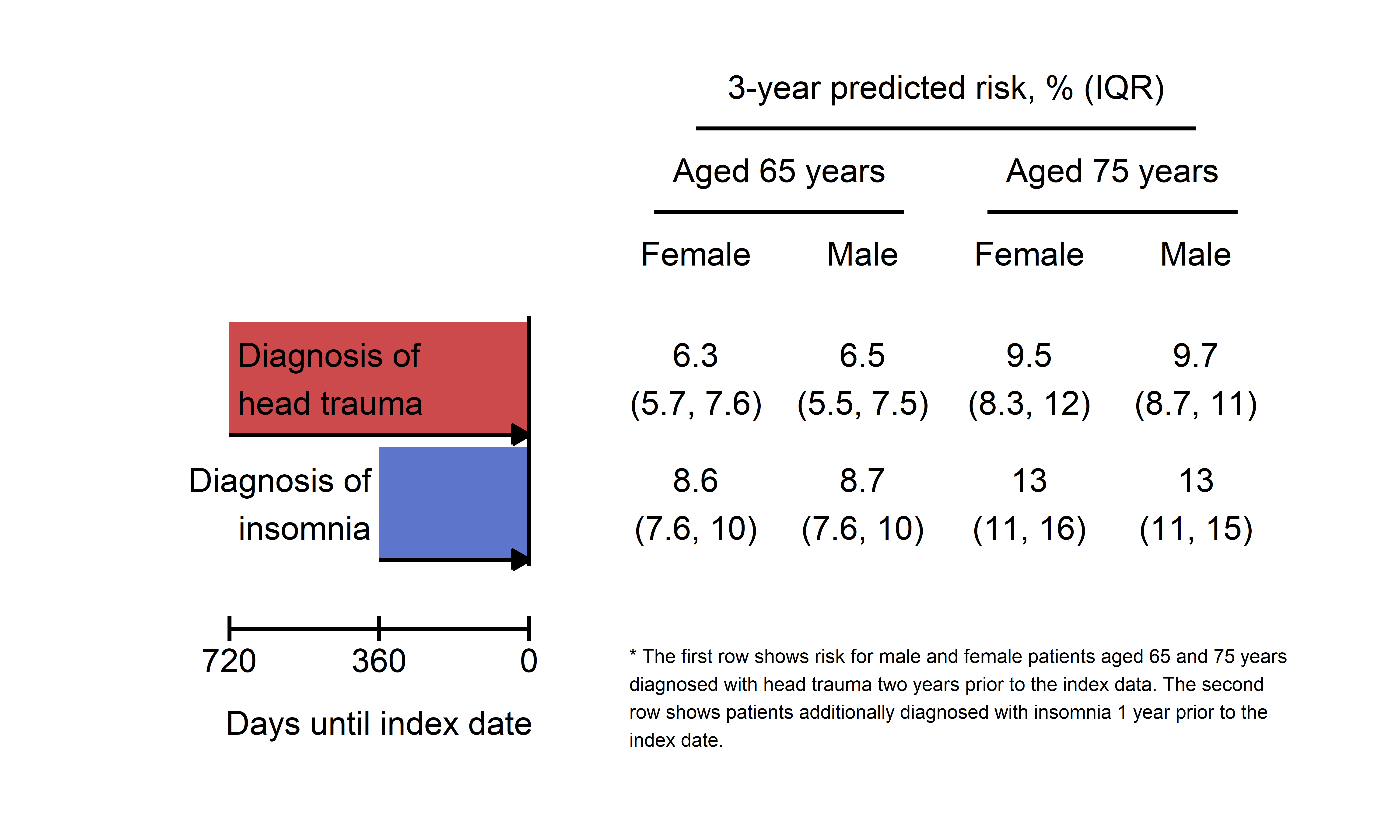

Figure S6: Multivariable adjusted 3-year predicted risk (25th, 75th percentile) for male and female patients aged 65 and 75 years. Prior to the index date, patients were diagnosed with hypertension (first row), hypertension and diabetes (second row), or hypertension, diabetes, and insomnia (third row).

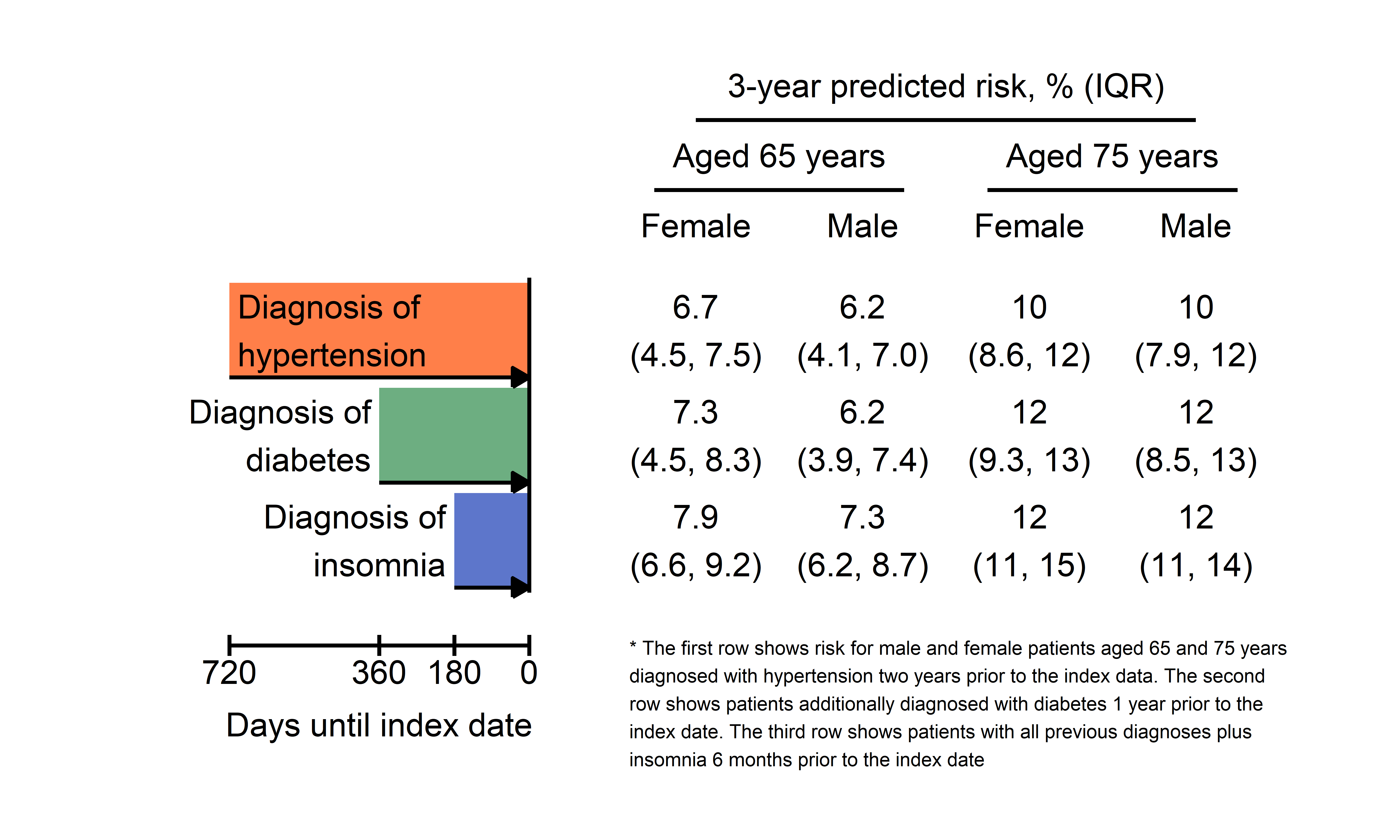
